# Beyond severity: Mapping cognitive heterogeneity in schizophrenia at the structural level

**DOI:** 10.64898/2026.02.25.26347062

**Authors:** Chen Chen

## Abstract

Cognitive heterogeneity is a fundamental aspect of schizophrenia (SCZ), but conventional domain-specific score-based approaches reduce cognition to single values, overlooking its structure and primarily reflecting the severity of cognitive impairment rather than qualitative differences. To address this gap, this study derived and validated a normative latent cognitive structure (N-LCS) serve as a reference to evaluate whether individual cognition aligns with the norm, providing a basis for characterizing cognitive heterogeneity in SCZ. Three deviation metrics represent the deviation magnitude (CDM), the angular deviation (CDA), and the domain driving the deviation (CDD). CDM showed a linear increase from healthy controls (HC) to unaffected siblings (UC) and SCZ patients. CDA differed between HC and the other two groups, but not between UC and SCZ. Reasoning remained the most frequent CDD, whereas processing speed declined across the SCZ liability spectrum. These findings imply that heterogeneous changes in cognitive structure already emerge in the UC stage, preceding both clinical onset and measurable score-level differences. Cognitive heterogeneity in SCZ can be delineated by three deviation dimensions: 53.1% of patients had reasoning as the CDD, with moderate CDM and CDA, suggesting that their cognitive structure is preserved relative to the N-LCS, whereas 31.2% had processing speed as the CDD, with larger CDM and intermediate CDA, and higher HAMA scores. The N-LCS captures signal beyond conventional score-based measures, supporting its utility and informing individualized treatment.

## Introduction

Cognitive impairment is a core feature of schizophrenia (SCZ) and a determinant of functional outcome [1]. Its clinical presentation is heterogeneous, with patients differing in both severity and cognition profiles, complicating clinical characterization and limiting targeted interventions [2].

Two approaches have been proposed to address this heterogeneity. Normative modeling establishes a population-level distribution and views SCZ as a departure from it [3]. In contrast, clustering-based approaches seek to identify patient subgroups with distinct cognitive phenotypes [4]. The resulting clusters often differ in the magnitude of severity rather than representing qualitatively distinct subtypes [5, 6].

Empirical evidence to date appears to favor interpreting the quantitative nature of this heterogeneity. Neuroimaging studies indicate that SCZ is associated with altered topological organization of brain networks [7, 8], and these anatomical changes being patient-specific [9]. Current clinical and research practice assesses cognition using domain-specific or composite scores, reducing cognition to single numerical values. Despite evidence that cognition can be understood as a network of interacting systems, with interdependent domains organized within a broader latent cognitive structure [10].

Building on this perspective, the present study derived and validated a normative latent cognitive structure (N-LCS) to model the structure of cognition, using it as a reference to evaluate whether individual cognition aligns with the norm. This approach moves beyond conventional score-based measures, enabling a structure-informed assessment of cognition and providing a basis for characterizing cognitive heterogeneity at the individual level in SCZ. Deviations from the N-LCS were explored to determine whether they provide additional information on symptoms beyond that captured by score-based measures, with cognitive specificity in SCZ grounded in its association with clinical symptoms [11]. Because unaffected siblings of SCZ patients (UC) represent a genetic high-risk population [12], the study also investigated how such deviations evolve along the schizophrenia liability spectrum, spanning healthy controls (HC), UC, and SCZ groups.

## 2. Methods

### 2.1 Data

The primary dataset comprised patients with SCZ (n = 32), UC (n = 32), and HC (n = 33), originally reported by Xu and Xian (2023) and publicly available [13].

External validation of the N-LCS was performed using the Center of Biomedical Research Excellence (COBRE) schizophrenia dataset [14], accessed via the Collaborative Informatics and Neuroimaging Suite Data Exchange tool (COINS; http://coins.mrn.org/dx). The COBRE dataset included 90 HC and 78 SCZ participants, but only the HC sample was used for validation.

Both datasets contained multi-domain neurocognitive measures, as well as demographic and clinical variables. All data were fully anonymized, and the present analyses did not require additional ethical approval.

### 2.2 Neurocognitive measures

To ensure comparability, analyses focused on four cognitive domains common to both datasets: processing speed, attention, working memory (WM), and reasoning.

In the primary dataset, these domains were assessed using the Wisconsin Card Sorting Test (WCST), Digit Span Forward (DSF), Digit Span Backward (DSB), Stroop Word, Stroop Color, Stroop Interference, Trail Making Test A (TMTA), and Trail Making Test B (TMTB). Three WCST subscores were included: correct responses, perseverative errors (PE), and trials to complete the first category (1st category).

To reduce dimensionality and address multicollinearity, Stroop performance was summarized as a composite score (Stroop-comp). For the Trail Making Test, the difference between TMTB and TMTA (TMT-diff) was calculated to reduce shared variance attributable to basic processing speed. Scores indicating poorer performance were reverse-coded so that higher values consistently indicated better cognitive performance.

In the COBRE dataset, cognitive measures were obtained from the MATRICS Consensus Cognitive Battery (MCCB) [15, 16], which provides standardized T-scores for the corresponding domains.

### 2.3 Data preprocessing

Data were screened for missing values and outliers prior to analysis. Cognitive variables were examined for skewness within the HC group and transformed when necessary, using order-normalization estimated in HC data and then applied to all participants.

In the primary dataset, cognitive variables were standardized using HC-based z-scores, with scores in the UC and SCZ groups expressed relative to the HC normative reference.

### 2.4 N-LCS and cognitive deviation measures

Parallel analysis in the HC group of the primary dataset suggested a single-factor solution [17]. Confirmatory factor analysis supported this solution (p = 0.137), although overall fit was modest (RMSEA = 0.10, 90% CI [0.00, 0.19]) [18]. A single-factor exploratory factor analysis was then used to derive the N-LCS, which served as the reference structure for subsequent analyses. The individual cognition was projected onto the N-LCS to estimate deviations from it.

In the COBRE dataset, the number of factors was determined using the Kaiser criterion [19], parallel analysis, and the minimum average partial (MAP) procedure [20]. Factor loadings were compared with those from the primary dataset to assess consistency.

Three measures were defined to characterize deviation from the N-LCS: cognitive deviation magnitude (CDM), cognitive deviation angle (CDA), and the cognitive deviation driver (CDD). CDM quantifies the magnitude of deviation from the N-LCS and reflects reduced overall similarity to the normative reference. CDA captures the angle between an individual’s cognitive structure and the N-LCS and indicates reorganization of inter-domain relationships. CDD was defined as the cognitive domain driving the individual’s deviation from the N-LCS.

### 2.5 Statistical analysis

Continuous variables were analyzed using the Kruskal–Wallis test, followed by pairwise Wilcoxon rank-sum tests with false discovery rate (FDR) correction. Correlations were evaluated using Spearman’s rank correlation coefficient. Associations between cognition and symptoms were examined via linear regression, with bootstrap resampling performed to assess the robustness of the estimates. Statistical significance was set at p < 0.05 (two-tailed). All analyses were conducted in R (version 4.5.0).

## 3. Results

### 3.1 Demographic characteristics

In the primary dataset, the three groups (HC, UC, SCZ) did not differ significantly in age, sex, or years of education (all p > 0.05), consistent with the original report [13]. Detailed demographic, cognitive, and clinical information is reported in the original study.

In the COBRE dataset, the HC group had a mean age of 37.5 years (SD = 11.8), 23 were female (25.6%), and the mean years of education was 13.8 (SD = 1.70).

### 3.2 External validation of N-LCS in COBRE dataset

Validation in the COBRE HC sample supported the stability of the N-LCS. The Kaiser criterion identified a one-factor solution, with the first eigenvalue (2.02) above 1 and subsequent values below [19]. Parallel analysis corroborated this solution, and the MAP selected one factor as optimal (Figure 1).

**Figure 1.**
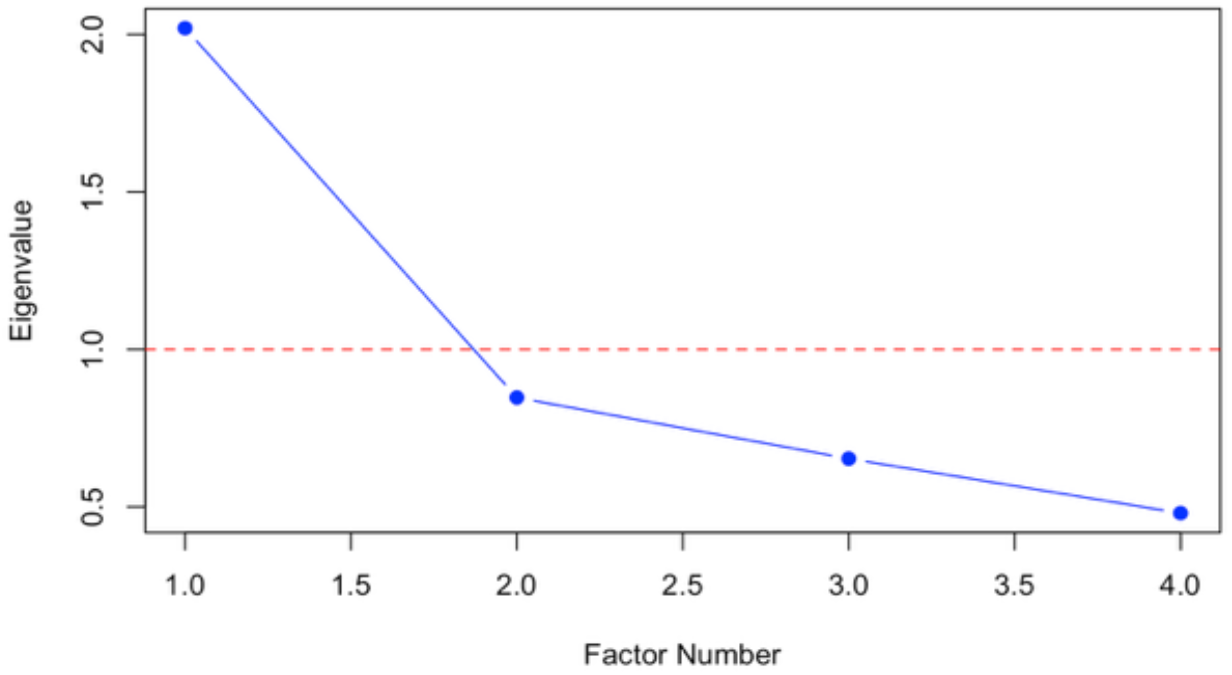
Scree plot of eigenvalues from parallel analysis in the COBRE HC sample

The factor loading pattern was consistent with that observed in the primary dataset, with stronger loadings for WM and attention and a weaker loading for reasoning.

### 3.3 N-LCS and score-based approach: comparative associations with negative symptoms

The N-LCS model revealed a significant association between CDA and negative symptom (β = -0.102, p = 0.005), which remained at a trend level after FDR correction (p = 0.07). Bootstrap analysis supported the stability of this effect, with a 95% confidence interval [-0.203, -0.047] and 99.9% of resamples in same direction. By contrast, the conventional score-based model showed no significant associations with negative symptoms (all p > 0.17). The N-LCS model explained more variance than the score-based model (R^2^ = 0.242 vs 0.084).

### 3.4 Cognitive deviations along the schizophrenia liability spectrum

CDM showed significant group differences across HC, UC, and SCZ (p = 2.49 × 10^-10^; see Table 1). Wilcoxon pairwise comparisons confirmed differences between all groups (all p < 0.05), with SCZ exhibiting the largest CDM. A significant linear trend revealed increasing CDM across groups (p= 2.34 × 10^-11^).

**Table 1.** CDM in HC, UC, and SCZ.

| Group | CDM | p-value |
| --- | --- | --- |
| HC | 2.50 (2.20, 2.87) |  |
| UC | 4.00 (3.23, 5.35) | $2.49 \times 10^{-10}^{**}$ |
| SCZ | 5.09 (4.01, 6.83) |  |

Data are presented as median (Q1, Q3) and compared using the Kruskal–Wallis test.
\*\* $p < 0.01$

CDA values were centered around zero in HC, whereas those in UC and SCZ were positively shifted (p = 0.012; see Table 2). Post hoc Wilcoxon tests demonstrated higher CDA in both UC and SCZ compared to HC (both p < 0.05). CDA was higher in SCZ than in UC, but this difference was not statistically significant (p = 0.81).

**Table 2.** Group differences in CDA.

| Group | CDA | p-value |
| --- | --- | --- |
| HC | 0.00 (-10.51, 7.94) |  |
| UC | 10.77 (2.72, 16.10) | 0.012* |
| SCZ | 12.51 (4.10, 18.95) |  |

Data are presented as median (Q1, Q3) and compared using the Kruskal–Wallis test.
CDA values are expressed as the angle (degrees). All values are relative to the HC baseline; negatives denote direction, not effect.
\* $p < 0.05$

**Table 3.** CDD proportions across HC, UC, and SCZ.

| Driver domain | HC | UC | SCZ |
| --- | --- | --- | --- |
| Attention | 9.09% | 6.25% | 9.38% |
| Processing speed | 9.09% | 18.8% | 31.2% |
| Reasoning | 51.5% | 68.8% | 53.1% |
| Working memory | 30.3% | 6.25% | 6.25% |

Values are percentages of subjects assigned to each CDD

**Table 3.**
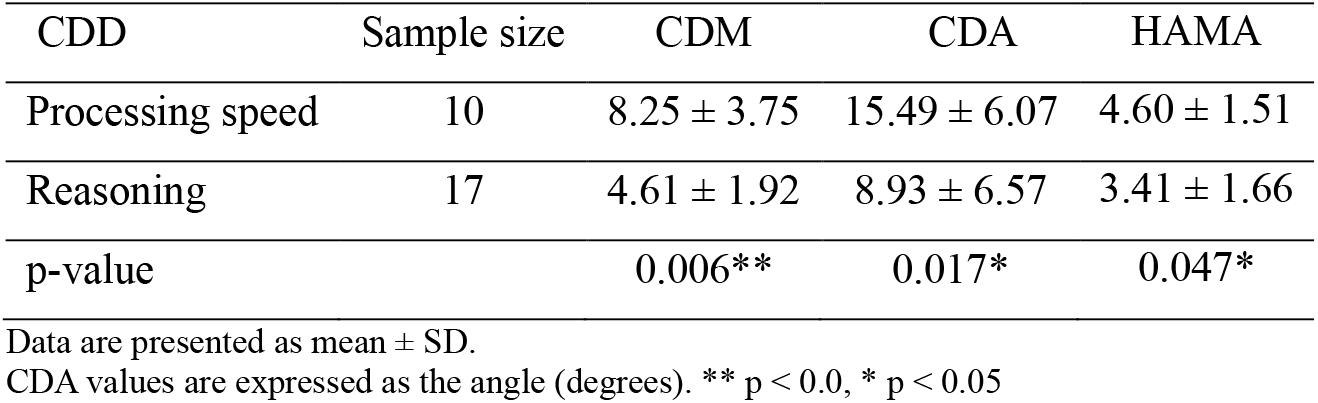
CDM and CDA across driver domains in the SCZ group.

| CDD | Sample size | CDM | CDA | HAMA |
| --- | --- | --- | --- | --- |
| Processing speed | 10 | $8.25 \pm 3.75$ | $15.49 \pm 6.07$ | $4.60 \pm 1.51$ |
| Reasoning | 17 | $4.61 \pm 1.92$ | $8.93 \pm 6.57$ | $3.41 \pm 1.66$ |
| p-value |  | 0.006** | 0.017* | 0.047* |

Data are presented as mean ± SD.
CDA values are expressed as the angle (degrees). \*\* $p < 0.0$ , \* $p < 0.05$

CDD distributions varied across groups (Table 3), although no statistical comparison was performed due to small cell counts in some categories. Reasoning remained the most frequent CDD in all groups, while processing speed became increasingly represented from HC to UC and SCZ, accompanied by a decrease in working memory.

### 3.4 Cognitive heterogeneity in SCZ

Cognitive heterogeneity in SCZ was delineated by CDM, CDA, and CDD. Patients with reasoning as the CDD had lower CDM and CDA than those with processing speed as the driver domain (Table 3). HAMA scores were lower in the reasoning group than in the processing speed group, while no significant differences were observed for other clinical symptoms (e.g., positive and negative symptoms). Sample sizes were small for attention (n=3) and WM (n=2), limiting interpretation for these domains. CDM and CDA were not significantly associated with disease course in SCZ (p = 0.36 and p = 0.57, respectively). The joint CDM and CDA distribution across all four driver domains is displayed in Figure 2 for visualization only.

**Figure 2.**
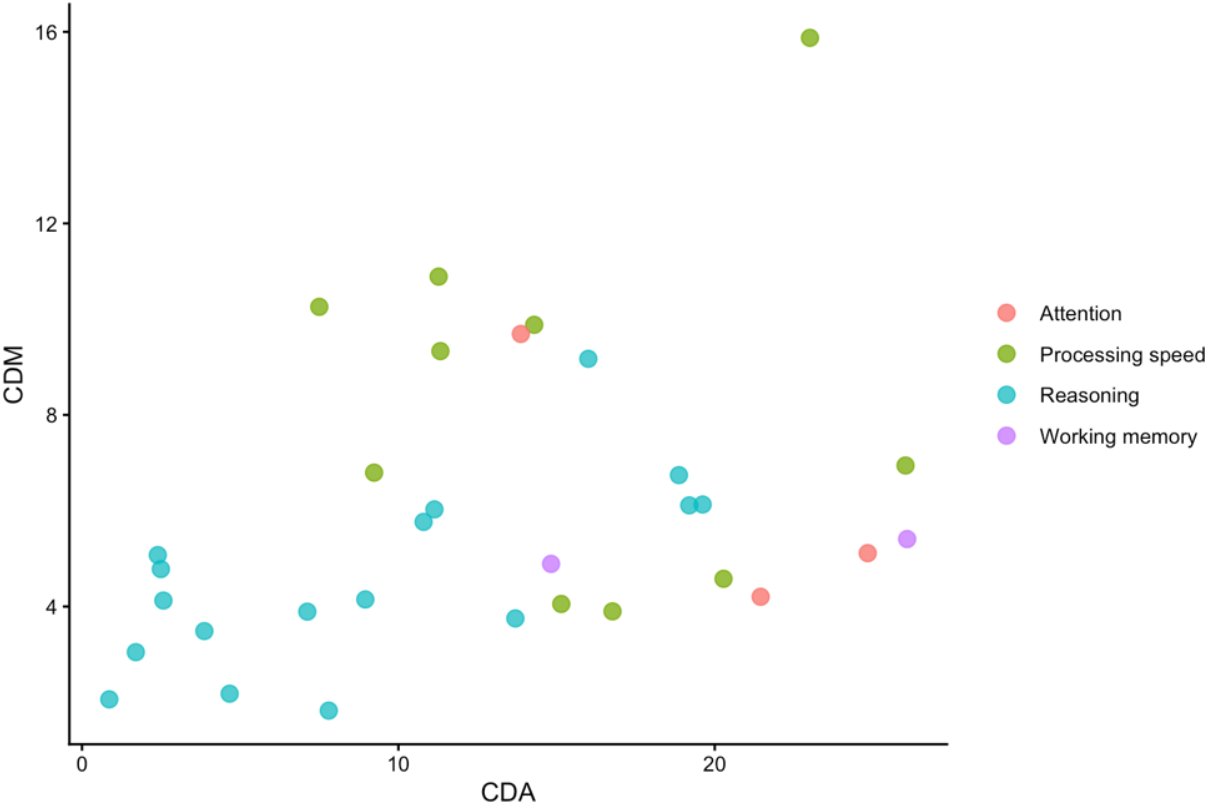
The joint distribution of CDM and CDA, colored by driver domain

## Discussion

A stable N-LCS captured the shared structure across cognitive domains, in line with evidence that cognitive performance is explained by a general cognitive factor [21-23]. Network neuroscience suggests that cognition mirrors connectivity within a network of distributed brain systems, with interdependent domains organized in a broader cognitive architecture that supports efficient information exchange and system-wide flexibility [10, 24].These perspectives provide a conceptual basis for the N-LCS. Both internal validation in the primary dataset and external validation in the COBRE dataset confirmed the stability of the N-LCS single-factor solution and its factor loadings, which remained consistent across demographic characteristics and cognitive test batteries [25].

In the primary dataset, the N-LCS model explained more variance than the score-based model, and CDA showed a significant association with negative symptom. One possible explanation is that negative symptom is not linked to deficits in any single cognitive domain [26], but rather to goal-directed, coordinated cognitive processes [27]. In this context, the N-LCS approach captures cognitive changes linked to clinical symptoms beyond those detected by score-based measures, supporting its suitability.

Based on the N-LCS, cognitive changes along the schizophrenia liability spectrum can be better characterized by the extent to which a person’s cognitive structure aligns with the norm. This view echoes the Hierarchical Taxonomy of Psychopathology (HiTOP) framework, which conceptualizes SCZ as an extreme manifestation along the dimensional continuum of psychopathology [28]. Altered inter-domain relationships, inferred from CDA, suggest potential cognitive structure reorganization that was already detectable at the UC stage and appeared comparable in SCZ [29, 30]. Nevertheless, the CDM-based assessment of overall dissimilarity from the N-LCS reveals a linear increase across groups [31], pointing toward the possibility that the strength of reorganized inter-domain relationships may continue to grow— a hypothesis that warrants further causal validation. At the domain level, this reorganization is not uniformly expressed but instead appears as a redistribution across domains. Reasoning remains the most frequent CDD, consistent with evidence that higher-order association cortices mature later than primary sensory regions [32], and may be more susceptible to disruption during development. Such vulnerability is also observed in resting-state studies of early psychosis, which have identified alterations in prefrontal and central executive networks [33]. Other domains, including WM and processing speed, exhibit observable trends across groups, but small cell sizes in some categories preclude further interpretation. In the same cohort, most neuropsychological test scores did not differ significantly between UC and SCZ [13], suggesting that cognitive reorganization may precede measurable score-level differences and potentially represent an early signal along the liability spectrum.

SCZ cognitive heterogeneity can be delineated in a three-dimensional map:CDM quantifies overall similarity to the N-LCS, CDA captures reorganization of inter-domain relationships, and CDD denotes the domain-level expression of deviations. Unlike conventional severity-based assessments [6, 34], this structure-informed approach integrates both quantitative and qualitative features, enabling a more comprehensive interpretation of cognitive heterogeneity at the individual level. In this cohort, patients with reasoning as the CDD exhibited limited structural deviation from the N-LCS. In contrast, patients with processing speed as the CDD showed larger deviations, consistent with evidence of reduced global efficiency with preserved local efficiency in functional brain networks, pointing to impaired integration across distributed systems [35]. The increasing proportion of processing speed as the CDD from UC to SCZ, despite similar genetic and environmental backgrounds, suggests that this subgroup may be sensitive to adversity and that processing speed may index this vulnerability at the domain level, providing a more specific expression of the broader reorganization described above. Clinically, HAMA scores differ across CDD-defined subgroups, with higher anxiety observed in processing speed group [36], highlighting that these subgroups may carry meaningful clinical differences rather than representing purely data-driven separation.

Several limitations should be acknowledged. First, the present analysis included four cognitive domains, which may not fully encompass the complexity of human cognitive structure. Future studies incorporating additional cognitive domains are needed to evaluate the robustness and generalizability of the N-LCS and its derived deviation metrics. Second, the cross-sectional design limits insights into the temporal dynamics of cognitive changes captured by the N-LCS and their potential prognostic implications. Third, the moderate sample size limited domain-level analyses for certain CDD categories and reduced statistical power to detect associations between N-LCS deviation metrics and clinical measures.

In conclusion, this study derived the stable N-LCS, a structure-informed approach that is sensitive to cognitive changes along the schizophrenia liability spectrum compared with conventional score-based measures. By characterizing individual-level cognitive heterogeneity in three dimensions, it goes beyond ‘intact-to-impaired’ outcomes. The N-LCS model shows significant associations with negative symptoms, which were not detected by score-based models, highlighting its potential clinical relevance. These findings support the utility of the N-LCS approach and suggest that further refinement and evaluation in larger and longitudinal cohorts could strengthen its application for characterizing cognitive heterogeneity in SCZ, with potential implications for individualized treatment.

## Data Availability

The primary data used in this study are publicly available from the original publication (Xu & Xian, 2023). The COBRE dataset was accessed via the Collaborative Informatics and Neuroimaging Suite Data Exchange tool (COINS; http://coins.mrn.org/dx).

## Acknowledgments

The primary dataset used in this study was obtained from Xu and Xian (2023). The validation dataset was downloaded from the Collaborative Informatics and Neuroimaging Suite Data Exchange tool (COINS; http://coins.mrn.org/dx). Data collection for the COBRE dataset was performed at the Mind Research Network and supported by a Center of Biomedical Research Excellence (COBRE) grant (5P20RR021938/P20GM103472) from the National Institutes of Health to Dr. Vince Calhoun.

The investigators associated with these datasets contributed to the design of the original research and data collection but did not participate in the analyses reported in this manuscript or in the preparation of this manuscript. The author gratefully thanks them for making these datasets publicly available.

